# Exploring the Genetic and Genomic Connection Underlying Neurodegeneration with Brain Iron Accumulation and the Risk for Parkinson’s Disease

**DOI:** 10.1101/2022.09.28.22280461

**Authors:** Pilar Álvarez Jerez, Jose Luis Alcantud, Lucia de los Reyes-Ramírez, Anni Moore, Clara Ruz, Francisco Vives Montero, Noela Rodriguez-Losada, Prabhjyot Saini, Ziv Gan-Or, Chelsea Alvarado, Mary B. Makarious, Kimberley J. Billingsley, Cornelis Blauwendraat, Alastair J Noyce, Andrew Singleton, Raquel Duran, Sara Bandres-Ciga

## Abstract

**Background:** Neurodegeneration with brain iron accumulation (NBIA) represents a group of neurodegenerative disorders characterized by abnormal iron accumulation and the presence of axonal spheroids in the brain. In Parkinson’s Disease (PD), iron accumulation is a cardinal feature of degenerating regions in the brain and seems to be a key player in mechanisms that precipitate cell death.

**Objectives:** The aim of this present study was to comprehensively explore the genetic and genomic connection between NBIA and PD etiology.

**Methods:** We screened the presence of known and rare pathogenic mutations in autosomal dominant and recessive genes linked to NBIA in a total of 4,481 PD cases and 10,253 controls from the Accelerating Medicines Partnership Parkinsons’ Disease Program and the UKBiobank. We further examined whether a genetic burden of NBIA variants contributes to PD risk through single-gene, gene-set, and single-variant association analyses. To investigate the potential effect of NBIA gene expression on PD, we assessed publicly available expression quantitative trait loci (eQTL) data through Summary-based Mendelian Randomization and conducted transcriptomic analyses in blood of 1,886 PD cases and 1,285 controls.

**Results:** Out of 28 previously reported NBIA screened coding variants, four missense were found to be associated with PD risk at a nominal p value < 0.05 (p.T402M-*ATP13A2*, p.T207M-*FA2H*, p.P60L-*C19orf12*, p.C422S-*PANK2*). No enrichment of heterozygous variants in NBIA-related genes risk was identified in PD cases versus controls. Burden analyses did not reveal a cumulative effect of rare NBIA genetic variation on PD risk. Transcriptomic analyses suggested that *DCAF17* is differentially expressed in blood from PD cases and controls.

**Conclusions:** Taking into account the very low mutation occurrence in the datasets and the lack of replication, our analyses suggest that NBIA and PD may be separate molecular entities, supporting the notion that the mechanisms underpinning iron accumulation in PD are likely not shared with NBIA. Elevated nigral iron levels may not contribute to PD etiology and may vary with anti-parkinsonian drugs used for treatment, environmental factors, or iron sequestration in tissue as a response to PD pathological change.

## Introduction

Neurodegeneration with brain iron accumulation (NBIA) represents a group of inherited heterogeneous neurodegenerative disorders characterized by iron accumulation and the presence of axonal spheroids in the basal ganglia and other brain areas.^1^ The worldwide prevalence of NBIA in the general population is estimated between 1-3/1,000,000 individuals, adding these disorders to the group of ultra-rare orphan diseases.^2^ In view of the rarity of these disorders and its diverse clinical presentation, NBIA can often go unrecognized, underdiagnosed, or misdiagnosed.

In the brain, the substantia nigra, putamen, globus pallidus and caudate nucleus have the highest iron concentration, and total iron content increases with age.^3^ Brain iron is crucial for important processes such as the synthesis of myelin and neurotransmitters and oxygen transport.^3,4^ Higher nigral iron concentrations have been shown in Parkinson’s Disease (PD) patients in postmortem studies repeatedly.^5,6^ This excess of intracellular iron is associated with oxidative stress, lipid peroxidation, cellular dysfunction, and neuronal death in PD and thus potentially playing a role in PD pathogenesis.^7,8^

To date, mutations in 13 genes have been associated with autosomal dominant (*FTL*), autosomal recessive (*ATP13A2, PANK2, PLA2G6, FA2H, CP, C19orf12, COASY, GTPBP2, DCAF17, VAC14*) and X-linked forms of NBIA (*WDR45, RAB39B*), and are involved in a wide range of molecular processes affecting mitochondrial function, coenzyme A metabolism, lipid metabolism and autophagy.^9^ Typically, NBIA disorders present with variable and complex phenotypes including parkinsonism, dystonia, intellectual disability, and cognitive decline.^1^ The NBIA clinical spectrum is broad and there is increased awareness of clinical overlap between different NBIA disorders as well as with other diseases such as Parkinson’s disease (PD). In PD, it has been widely suggested that genetic components contributing to disease might include rare genetic variants of small or moderate effect, where functional and deleterious alleles might exist.^10^ Furthermore, it has been previously predicted that variability in genes causing young-onset autosomal recessive neurological diseases contribute to late-onset neurological diseases. In fact, homozygous and compound heterozygous loss-of-function mutations in *TREM2* have been previously associated with an autosomal recessive form of early-onset dementia, while when found in heterozygous state confer moderate risk for Alzheimer’s disease.^11,12^ Similarly, homozygous and compound heterozygous mutations in *GBA* are responsible for Gaucher’s disease while heterozygous variants are a well-validated risk factor for PD.^13^

In PD, iron accumulation is a cardinal feature of degenerating regions in the brain and has been implicated in mechanisms that precipitate cell death.^14^ Iron is a transition metal involved in several cellular functions such as neuronal metabolism, DNA synthesis, oxygen transport, mitochondrial respiration and myelination in the brain.^15^ Furthermore, it is implicated in production and turnover of some neurotransmitters such as dopamine, epinephrine or serotonin.^16^ Iron tends to accumulate with age mainly in the cortex and the nuclei of the basal ganglia, including the globus pallidus, putamen and caudate nucleus; and an excessive accumulation in these regions is associated with neurodegenerative disorders.^17^ Evidence suggests abnormal iron levels in the brains of PD patients and a role for iron dysregulation in the disease.^18^ Interestingly, NBIA-related genes are involved in mitochondrial function and autophagy, dysfunction of which is implicated in PD, suggesting there could be shared molecular pathways and gene networks between NBIA and PD.^19^ Furthermore, autopsy examination of NBIA-genetically confirmed cases has demonstrated Lewy bodies in some subforms, linking the pathology of both diseases.^20^

The aim of the present study was to explore the relationship between genes known to contain mutations that cause autosomal dominant/recessive NBIA and PD etiology. Using the largest genetic and genomic datasets of PD cases and controls to date including the Accelerating Medicines Partnership Program for Parkinson’s Disease (AMP-PD) and the UKBiobank (UKB), we screened for the presence of known and rare pathogenic mutations linked to NBIA in PD. We further examined whether a genetic burden of variants in NBIA-linked genes could contribute to the risk of developing PD by performing single gene, gene-set, and single variant association analyses. Finally, to investigate the potential effect of changes in NBIA-linked genes expression in PD compared to healthy individuals, we assessed publicly available expression quantitative trait loci (eQTL) results from GTEx v8, BRAINEAC and transcriptomics data from AMP-PD.

## Methods

### Whole-genome sequencing data

Whole-genome sequencing (WGS) data was obtained from the Accelerating Medicines Partnership - Parkinson’s Disease Initiative release 2.5 (AMP-PD; www.amp-pd.org) which contained 3,376 PD patients and 4,610 healthy unrelated controls of European ancestry, with an average age at onset of 62 years in cases and age at collection of 72 years in controls. Detailed cohort characteristics, as well as quality control procedures, are further described in https://amp-pd.org/whole-genome-data. Data generation is described in detail by Iwaki and colleagues.^21^ NBIA related variants were extracted from the data by using PLINK1.9 and annotated with ANNOVAR. Association analyses of NBIA variants and risk for PD were performed using logistic regression under RVTESTS package v2.1.0 default parameters and adjusted by sex, age, and at least 5 PCs to account for population substructure.^22^

To assess the cumulative effect of multiple rare variants on the risk for PD, we performed single gene and gene-set burden analyses. Including all variants within the gene boundaries, a minimum allele count (MAC) threshold of 1 was applied. Sequence Kernel Association Test (SKAT-O) was performed considering two variant frequency levels: MAF < 1 % and MAF < 3%.. Age, sex, and at least 5 PCs, were accounted for as covariates.

### Whole exome sequencing data

Whole-exome sequencing (WES) data was obtained from the UK Biobank 2021 release (UK Biobank; https://www.ukbiobank.ac.uk/) which contained 1,105 PD patients and 5,643 healthy unrelated controls of European ancestry, with an average age at recruitment of 63 years in cases and 64 years in controls. Detailed cohort characteristics, as well as quality control procedures, are further described in https://www.ukbiobank.ac.uk/enable-your-research/about-our-data/genetic-data. NBIA related variants were extracted from the data by using PLINK1.9 and annotated with ANNOVAR.^23,24^ Association analyses of NBIA variants and risk for PD were performed using logistic regression in RVTESTS package v2.1.0 adjusted by sex, age, and at least 5 PCs to account for population substructure.^22^ Single gene and gene-set burden analyses were performed mimicking the AMP-PD pipeline mentioned above.

### Transcriptomics data

Whole blood time progression gene expression data (gencode v29) from 0-24 months after first study visit was accessed from AMP-PD. We used PPMI (https://amp-pd.org/unified-cohorts/ppmi), PDBP (https://amp-pd.org/unified-cohorts/pdbp), and BioFIND (https://amp-pd.org/unified-cohorts/biofind) cohorts at the baseline of the study, 0 month time point (note BioFIND ‘Baseline’ is at ‘0.5’ month), including a total of 1,886 cases and 1,285 control samples. Expression data were quantified as transcripts per kilobase million (TPM) and quantile normalized before NBIA related genes values were extracted using Ensembl gene ids. Using the scikit-learn Python packages 19, residuals were calculated using linear regression, and the data were adjusted using age, sex, race, at least 5 PCs, cohort, and missingness rate as covariates. To test for significant differences in expression between the cases and controls, we performed a t-test with the residuals. We also used an additional dataset taken from the 24-month time point in the PDBP and PPMI cohorts of 841 cases and 386 controls to confirm our findings.

### Summary based Mendelian Randomization

We use Summary data-based Mendelian Randomization (SMR) as a complementary method to further explore putative associations between gene expression levels in NBIA genes and PD risk. SMR is a mendelian randomization (MR) method that uses summary-level data to test if an exposure variable (i.e., gene expression) and outcome (i.e., trait) are associated because of a shared causal variant. In order to distinguish pleiotropy from linkage, the heterogeneity in dependent instruments (HEIDI) method was applied to each tested single nucleotide variant (SNV).^25^ SMR and HEIDI analysis were conducted using the SMR package.^25^

In order to conduct the analysis, we used PD GWAS summary statistics as well as methylation and eQTL meta-analysis summary statistics.^26,27^ PD GWAS summary statistics from Nalls et al. 2019 contained 17 datasets which consisted of 37,688 PD cases, 18,618 proxy cases, and 1,417,791 controls.^28^ The methylation data we used consists of brain and blood gene expression data from a meta-analysis conducted by Qi et al. 2018. All eQTL analyses were performed to test for potential in cis association between methylation site and SNV. In order to achieve this, SNVs within 1Mb of the probes of interest were selected.^26^ The eQTL summary statistic data used were generated from peripheral blood and came from the meta-analysis conducted by Wu et al 2018. All of the SNPS from Wu et al. are located within 2Mb of a probe.^29^

In order to prioritize SNP candidates, results were initially filtered by chromosome and base pair position for the *DCAF17* gene in hg19. Afterwards, the resulting candidates were filtered by the SMR multi p-value at a threshold of p < 0.05.

## Results

### Genetic screening of NBIA related genes in whole genome and whole exome sequencing data of Parkinson’s disease cases and controls

#### ATP13A2

Genetic variants in the *ATPase Cation Transporting 13A2* (*ATP13A2*) gene, located on chromosome 1, have been previously associated with Kufor-Rakeb syndrome, spastic paraplegia type 78, and parkinsonism.^30,31,32^ A total of 36 protein-coding variants were present in the screened datasets, of which four missense variants had a higher frequency in cases versus controls. All variants found in the studied genes are listed in Sup. Table 1. Out of these four variants, three were present in the AMP-PD cohort and one in UKB exome data. The variants found in AMP-PD (p.I946F, p.A249V, p.T12M) have been previously associated with PD in individuals of European ancestry although their pathogenicity is still unclear.^33,34^ Of note, p.T12M was found in heterozygous state in a patient with Young Onset Parkinson’s disease (YOPD) without atypical symptoms and no other mutations in known PD genes.^34^ Further study into this mutation has shown that it does not seriously alter protein stability but that it may impair ATPase activity.^35^ The p.T12M mutation was present in our data in one male PD case with an age at inclusion of 72 years old and absent in controls. The p.T402M variant has been previously described in an individual with ataxia-myoclonus syndrome with an age at onset of 38 and lacking parkinsonism.^35,36^ The report postulates that together, progressive ataxia with action myoclonus along with lack of parkinsonism may suggest a new phenotype linked to *ATP13A2*. In the UKB cohort, a heterozygous male carrier of the p.T402M mutation with age at recruitment of 55 years had diagnosis of PD. This mutation was absent in controls.

**Table 1:** Results of transcriptomic analysis using blood PPMI data. *DCAF17* is the only NBIA gene with significant differential expression in PD.

| Gene | Ensemble ID | T-statistic | P-value |
| --- | --- | --- | --- |
| ATP13A2 | ENSG00000159363.17 | -0.5126 | 0.6083 |
| DCAF17 | ENSG00000115827.13 | -4.036 | 5.537x10<sup>-5</sup> |
| GTPBP2 | ENSG00000172432.18 | -0.3596 | 0.7192 |
| VAC14 | ENSG00000103043.14 | -0.5266 | 0.5985 |
| COASY | ENSG00000068120.14 | 0.4632 | 0.6432 |
| C19orf12 | ENSG00000131943.17 | -1.446 | 0.1482 |
| FTL | ENSG00000087086.14 | 0.655 | 0.5125 |
| PANK2 | ENSG00000125779.22 | 0.8218 | 0.4113 |

#### CP

The *Ceruloplasmin (CP*) gene, located on chromosome 3, is closely related to iron metabolism in the brain.^37^ A total of 29 coding variants were present in our datasets, of which four distinct missense variants had a higher frequency in cases than controls. Notably, two of these variants, p.R793H and p.P477L, were seen in both AMP-PD and UKB. There have been reports linking lower *CP* levels with PD development, and *CP* gene mutations have been associated with substantia nigra hyperechogenicity using transcranial sonography.^38^ Additionally, hyperechogenicity is a common finding in idiopathic PD patients and many studies suggest that it is an important risk marker of future PD.^39,40^ One mutation associated with SN hyperechogenicity is p.R793H.^41^ The original study identified eight individuals of German origin carrying this variation, out of which five were PD patients and three were controls. All but one of the controls showed increased substantia nigra hyperechogenicity. Our study identified the p.R793H variant in homozygosity in one case and one control in the AMP-PD cohort, while it was absent in cases and present in two controls in UKB. Additionally, we found p.R793H in heterozygosity in 53 cases and 46 controls (F_A=1.11×10^−2^, F_U=7.78×10^−3^) in the AMP-PD cohort and in 28 cases and 153 controls (F_A=1.27×10^−2^, F_U=1.23×10^−2^) in UKB. However, it did not reach statistical significance for either dataset. Of note, we observed biallelic heterozygosity in one case and two controls from UKB carrying both the p.P477L and p.R793H mutations. The p.D544E variant, also reported to be associated with substantia nigra hyperechogenicity in parkinsonism, was only found in UKB.^41^ The p.I329F variant was only present in UKB data. A three base pair deletion at this position has been associated with aceruloplasminemia, a disorder in which iron gradually accumulates in the brain and other organs.^42^ Additionally, this is a multi-allelic variant for which the risk allele identified in our analyses differs from those reported on The Human Gene Mutation Database (HGMD).

#### FA2H

In the *Fatty Acid 2-Hydroxylse* (*FA2H*) gene, located on chromosome 16, we found 28 coding variants out of which two missense variants had a higher frequency in cases. The variant p.E78K appeared in both datasets and is associated with the fatty acid hydroxylase-associated neurodegeneration(FAHN)/hereditary spastic paraplegia (SPG35) phenotype.^43^ The FAHN/SPG35 phenotype manifests with early childhood onset predominantly lower limb spastic tetraparesis and truncal instability, dysarthria, dysphagia, cerebellar ataxia, and cognitive deficits, often accompanied by exotropia and movement disorders.^43^ At the time of the first report, p.E78K was labeled as very rare, MAF < 0.01, and it was found in two individuals from different families of European ancestry, however no conclusions were drawn about its significance. The two patients presented with gait variation at age 20 and 14 and later became wheelchair dependent after a median disease length of seven years. The p.E78K mutation has also been reported in a patient with an age at onset of 10 with a positive family history of spastic paraplegia and was labeled as a putative pathogenic variant.^44^ In AMP-PD, two cases and two controls (F_A=4.05×10^−4^, F_U=3.24×10^−4^) along with one case and five controls (F_A=4.53×10^−4^, F_U=4.43×10^−4^) in UKB were heterozygous carriers of the p.E78K mutation. The PD carriers under study with the p.E78K mutation had an age of inclusion of 70 and 52 for AMP-PD and 69 for UKB. The case/control frequency for p.E78K did not reach significance in our analysis. For the p.T207M mutation, all previously reported carriers were compound heterozygous with additional mutations in *FA2H* and had the spastic paraplegia phenotype.^31,43,45^ One of these carriers had a healthy mother with the p.T207M mutation in heterozygosity. In our study, we found one male UKB case with an age at inclusion of 65 carrying the p.T207M mutation. This mutation was absent in controls and was the only *FA2H* mutation that reached statistical significance.

#### C19orf12

Variation in the *Chromosome 19 open reading frame 12* (*C19orf12*) gene, located on chromosome 19, has been associated with the mitochondrial membrane protein-associated neurodegeneration (MPAN) subtype of NBIA which presents with cognitive decline progressing to dementia, prominent neuropsychiatric abnormalities, motor neuronopathy, and parkinsonism movement abnormalities.^46,47,48^ The earliest and most common symptom is gait abnormality marked by an early age at onset (4-30 years old). Out of 18 coding variants present, four missense variants in our datasets showed a higher frequency in cases versus controls, with only one proving significant association and previously linked to the MPAN phenotype in the literature. In AMP-PD data, we found two multiallelic variants, p.G65V present in one case and one control and p.G65A present in just one case and no controls. In two patients reported in the literature, compound heterozygosity for p.G65V and p.G69RfsX10 variants led to the MPAN phenotype.^47,48^ Additionally, another two patients were reported to have this phenotype; one carrying the compound heterozygous mutations p.G65V and p.P60L and the other carrying a homozygous p.G65V variant.^49^ In UKB data, we found three cases and 11 controls (F_A=1.36×10^−3^, F_U=9.75×10^−4^) with the variant p.K142E. In the literature, this variant has been seen in compound heterozygosity in patients with the MPAN NBIA phenotype.^50,51,52^ Lastly, we found the p.P60L mutation at statistical significance present in one UKB case and absent in controls. The carrier under study was a female PD patient with an age at inclusion of 66 and no other notable *C19orf12* mutations. The p.P60L variant has been previously detected in an MPAN patient who also carried the p.G65E *C19orf12* mutation.^48^

#### FTL

Our study identified only one missense variant (p.E104) out of nine with a higher frequency in cases than controls in the *Ferritin Light Chain* (*FTL*) gene, located on chromosome 19. The p.E104 mutation was only of note in the AMP-PD dataset and was present in one case and one control (F_A=2.02×10^−4^, F_U=1.62×10^−4^). The PD case under study was a male with an age at inclusion of 68 years old. The p.E104 variant has been reported as causing an L-ferritin deficiency phenotype.^48,53^ This variant has been previously described in a homozygous state in an individual of Italian ancestry with an age at onset of seven years old. The patient was affected by idiopathic generalized seizures during infancy, atypical restless leg syndrome, and some mild neuropsychological impairment. This was the first report of a complete loss of *FTL* function.

#### PANK2

Variation in the *Pantothenate Kinase 2* (*PANK2*) gene, located on chromosome 20, has been previously associated with pantothenate kinase-associated neurodegeneration (PKAN), the most common subtype of NBIA.^54^ Clinical symptoms include dystonia, dysarthria, muscular rigidity, poor balance, and spasticity.^54,55^ We found six distinct coding variants in our dataset, out of 68 present, with a higher frequency in cases. Two of these variants, one in AMP-PD (p.K406R) and one in UKB (p.C422S), were multiallelic and are listed on HGMD under different risk alleles (p.K406I and p.C422R respectively). Our remaining variants were composed of missense/nonsense mutations (p.T234A, p.G521R, p.D46V) and one small deletion (p.Leu425del). The p.G521R mutation was the only one present in both AMP-PD and UKB. This variant results in an unstable and inactive PanK2 protein and has been reported as the most common variant in *PANK2*.^56^ It is often found in a compound heterozygous state in individuals presenting with the PKAN phenotype.^56,57^ In AMP-PD, two PD cases, one male and one female with ages at inclusion of 54 and 67 respectively, carried the p.G521R mutation as compared to one control (F_A=4.05×10^−4^, F_U=4.05×10^−4^). In UKB, three cases, all males with an average age at inclusion of 62 years old, presented with the p.G521R mutation compared to five controls (F_A=1.36×10^−3^, F_U=4.43×10^−4^). No cases or controls from either dataset presented with other HGMD-classified *PANK2* mutations.

#### PLA2G6

In the *Phospholipase A2 Group VI* (*PLA2G6*) gene, located on chromosome 22, we found 100 variants of which seven missense variants had a higher frequency in cases versus controls. These variants have been previously associated with both infantile neuroaxonal dystrophy (INAD) and PD.^58,59,60,61,62^ INAD is a severe progressive psychomotor disorder, which presents before the third year of life, characterized by the presence of axonal spheroids throughout the central and peripheral nervous system.^62,63,64^ Most patients suffer neurological regression, following a period of normal development. The more common signs are oculomotor disorders, axial hypotonia, pyramidal signs, axonal neuropathy, gait impairment,optic atrophy, psychiatric symptoms and dysphagia between others. The variant p.M470V is the only variant out of three associated with the INAD phenotype that was present in both the AMP-PD and UKB cohorts.^65^ In AMP-PD, one female PD patient carried the p.M470V mutation with an age at inclusion of 73 years and one control (F_A=2.02×10^−4^, F_U=1.62×10^−4^). In UKB, two PD patients, both male, presented with this mutation with ages at inclusion of 62 and 50, respectively and 2 controls (F_A=9.05×10^−4^, F_U=1.77×10^−4^). Additionally, p.A343T has been associated with PD and we identified 77 cases and 86 controls carrying this mutation in AMP-PD (F_A=1.58×10^−2^, F_U=1.41×10^−20^) and 29 cases and 130 controls in UKB data (F_A=1.31×10^−2^, F_U=1.15×10^−2^).^59,65^ Of note, one of the p.A343T AMP-PD case carriers was a female homozygous for the mutation with an age at inclusion of 65. Additionally, a heterozygous carrier of p.A343T in the UKB was a PD case and additionally carried the p.S34L mutation, also associated with PD.^59,65^ This patient had an age at inclusion of 60. However, no *PLA2G6* mutation with a higher frequency in cases reached statistical significance.

### Transcriptomic analyses

Two genes including *FA2H* (ENSG00000103089.8) and *CP* (ENSG00000047457.13) were removed for further analyses based on poor call rates. Of the 10 NBIA genes left, *DCAF17* expression (ENSG00000115827.13) was found to be significantly different in cases versus controls (Figure 2)(Table 1). To further investigate these results we looked into SMR analysis for the *DCAF17* gene. To prioritize SNV candidates, results were initially filtered by chromosome and base pair position for the *DCAF17* gene in hg19. Afterwards, the resulting candidates were filtered by the SMR multi p-value at a threshold of p < 0.05. SMR analyses for the *DCAF17* gene revealed four eQTL signals with a significant SMR p-value. Out of these four eQTL signals, only one was significant in both SMR analyses (p = 0.0024) and Nalls et. al 2019 GWAS (p = 0.0004). A summary of the DCAF17 SMR data can be found in Supplementary Table 2.

**Figure 1:**
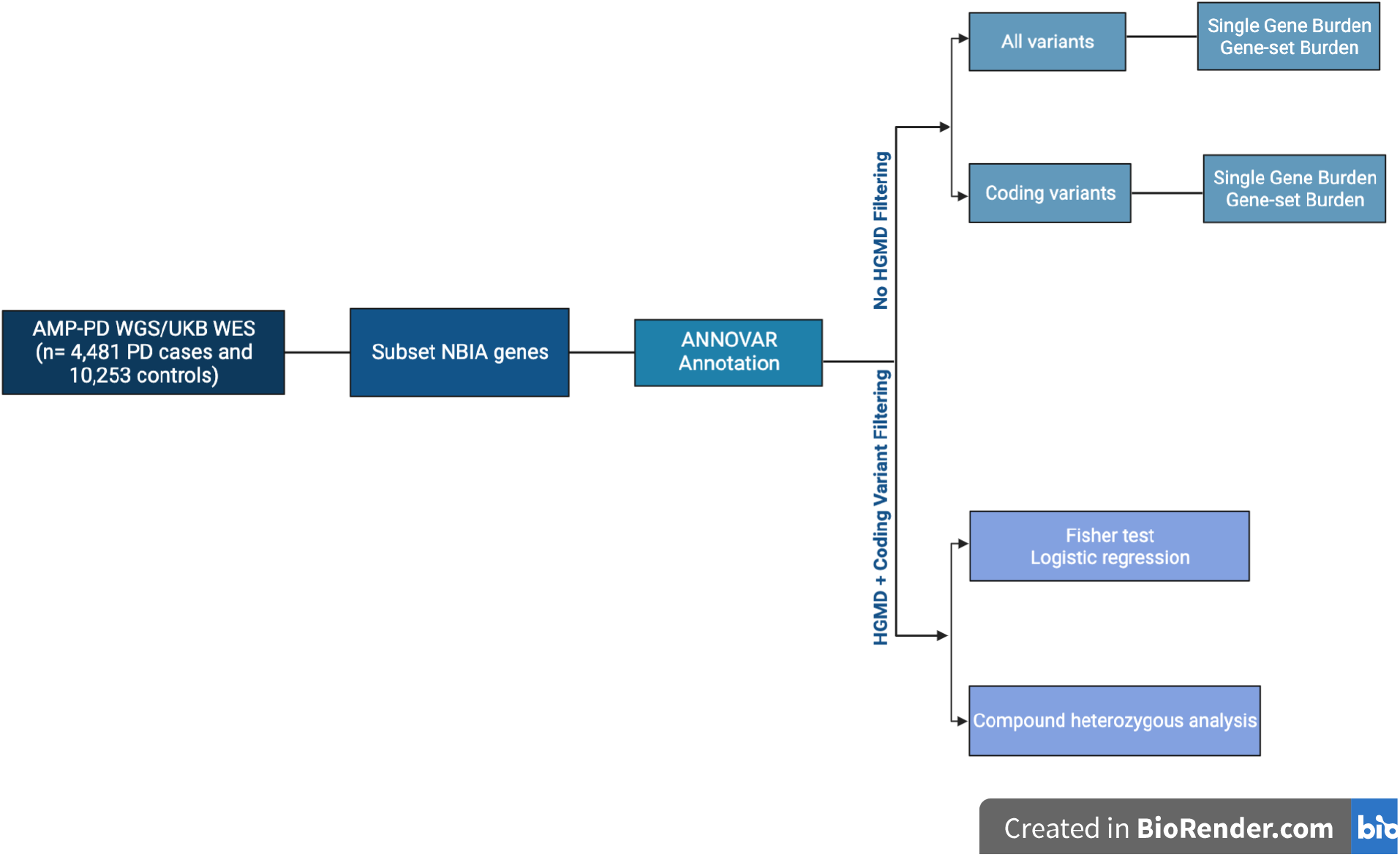
Overview of methods for association and burden analyses.

**Figure 2:**
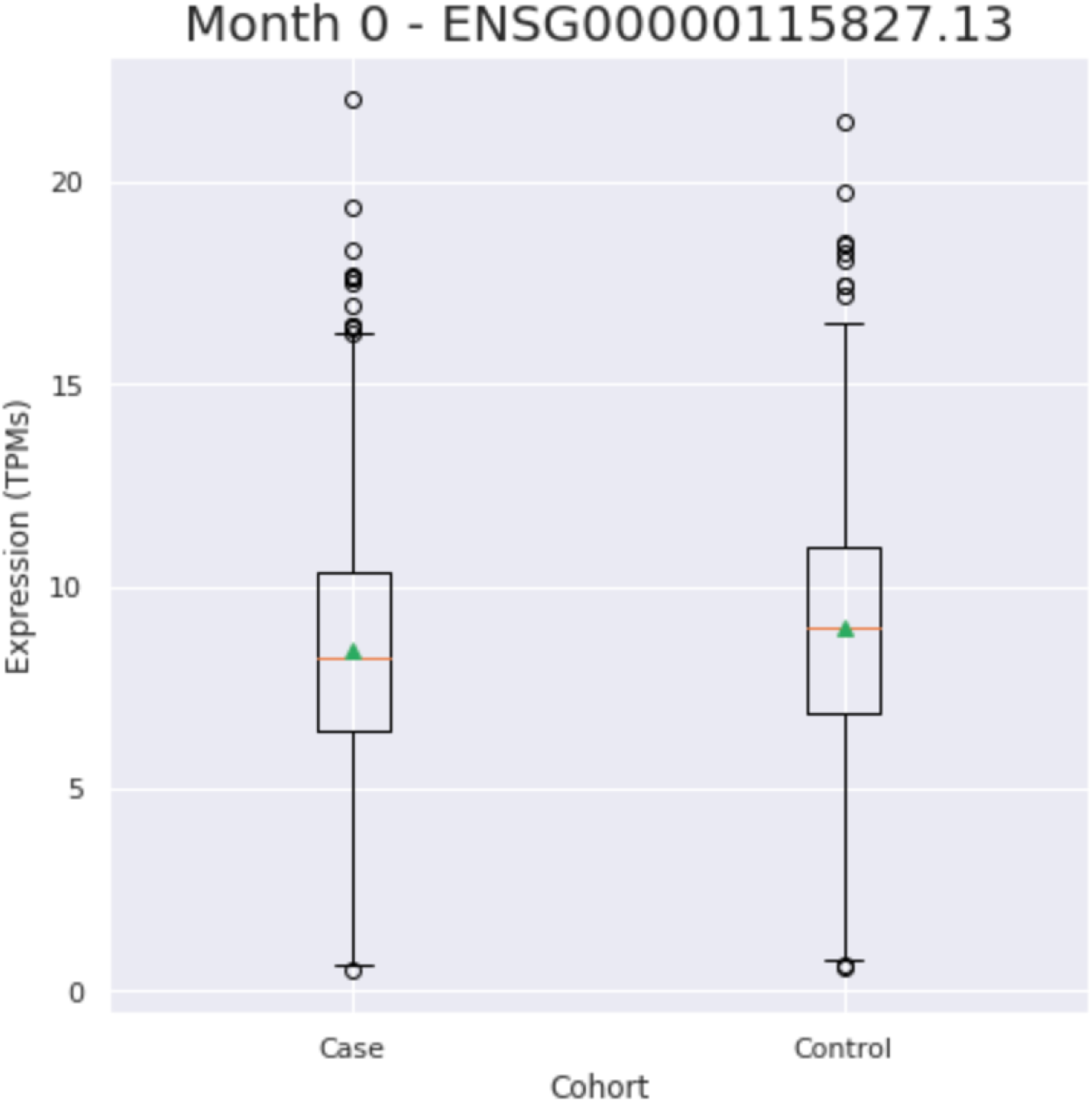
*DCAF17* expression in blood of PD cases versus controls.

### Cumulative effect of genetic variation through single gene and gene-set burden analyses

When considering the cumulative effect of rare variation on PD risk, single gene and gene-set burden analyses did not reveal any statistically significant differences between cases and controls at MAF < 0.01 or MAF < 0.03 in any of the three cohorts and after meta-analysis (Table 2).

**Table 2:** Results of burden analysis at two minor allele frequencies for NBIA genes in PD. MAF= Minor Allele Frequency, P_AMP= P value in AMP_PD cohort, P_UKB= P value in UKB cohort, P_Meta= P value in meta analysis, Weighted Z = Weighted Z Statistic.

| Gene | Variant Group | MAF | P_AMP | P_UKB | P_Meta | WeightedZ |
| --- | --- | --- | --- | --- | --- | --- |
| ATP13A2 | All | 0.01 | 0.0904 | 0.8813 | 0.2811 | 0.6518 |
| ATP13A2 | All | 0.03 | 0.6599 | 0.5547 | 0.7339 | -0.6615 |
| ATP13A2 | Coding | 0.01 | 0.9272 | 0.2949 | 0.6281 | -1.0192 |
| ATP13A2 | Coding | 0.03 | 0.6622 | 0.3033 | 0.5233 | -1.2470 |
| C19orf12 | All | 0.01 | 0.8789 | 0.5115 | 0.8089 | 1.6650 |
| C19orf12 | All | 0.03 | 0.7091 | 0.5115 | 0.7305 | 1.2089 |
| C19orf12 | Coding | 0.01 | 1.0000 | 0.4262 | 0.7897 | 1.6592 |
| C19orf12 | Coding | 0.03 | 1.0000 | 0.4262 | 0.7897 | 1.6592 |
| COASY | All | 0.01 | 0.5435 | 0.0832 | 0.1853 | -0.5970 |
| COASY | All | 0.03 | 0.5435 | 0.0832 | 0.1853 | -0.5970 |
| COASY | Coding | 0.01 | 1.0000 | 0.0936 | 0.3154 | 0.4017 |
| COASY | Coding | 0.03 | 1.0000 | 0.0936 | 0.3154 | 0.4017 |
| CP | All | 0.01 | 0.3392 | 0.4920 | 0.4657 | 1.0089 |
| CP | All | 0.03 | 0.6747 | 0.4339 | 0.6524 | 0.2643 |
| CP | Coding | 0.01 | 0.6615 | 0.3514 | 0.5717 | 0.4431 |
| CP | Coding | 0.03 | 0.6615 | 0.2169 | 0.4221 | 0.1493 |
| DCAF17 | All | 0.01 | 0.2382 | 0.2288 | 0.3721 | -0.9574 |
| DCAF17 | All | 0.03 | 0.2382 | 0.7715 | 0.8393 | 0.2144 |
| DCAF17 | Coding | 0.01 | 0.1146 | 0.4555 | 0.6522 | 0.8371 |
| DCAF17 | Coding | 0.03 | 0.1146 | 0.7161 | 0.8917 | 1.2486 |
| FA2H | All | 0.01 | 1.0000 | 0.3676 | 0.7355 | -0.4057 |
| FA2H | All | 0.03 | 0.8598 | 0.3676 | 0.6801 | -0.7388 |
| FA2H | Coding | 0.01 | 0.5816 | 0.5064 | 0.6546 | -0.9324 |
| FA2H | Coding | 0.03 | 0.5397 | 0.5064 | 0.6278 | -0.8240 |
| FTL | All | 0.01 | 0.4990 | 0.8460 | 0.7862 | -0.1314 |
| FTL | All | 0.03 | 0.4990 | 0.8460 | 0.7862 | -0.1314 |
| FTL | Coding | 0.01 | 0.5583 | NA | NA | 0.0555 |
| FTL | Coding | 0.03 | 0.5583 | NA | NA | 0.0555 |
| GTPBP2 | All | 0.01 | 0.5471 | 0.1980 | 0.3491 | -0.7209 |
| GTPBP2 | All | 0.03 | 0.2766 | 0.4241 | 0.3687 | -0.8013 |
| *GTPBP2* | Coding | 0.01 | 0.0696 | 0.3808 | 0.1227 | -1.6865 |
| *GTPBP2* | Coding | 0.03 | 0.6000 | 0.3712 | 0.5572 | 0.7064 |
| *PANK2* | All | 0.01 | 0.3261 | 0.8209 | 0.6205 | 1.5979 |
| *PANK2* | All | 0.03 | 0.4622 | 0.9000 | 0.7809 | 0.8418 |
| *PANK2* | Coding | 0.01 | 0.7875 | 0.0507 | 0.1684 | 0.3729 |
| *PANK2* | Coding | 0.03 | 0.7875 | 0.0507 | 0.1684 | 0.3729 |
| *PLA2G6* | All | 0.01 | 0.7644 | 0.2124 | 0.4575 | 1.3405 |
| *PLA2G6* | All | 0.03 | 0.7661 | 0.3815 | 0.6518 | 0.4891 |
| *PLA2G6* | Coding | 0.01 | 0.9007 | 0.3588 | 0.6882 | -0.9407 |
| *PLA2G6* | Coding | 0.03 | 0.8603 | 0.6690 | 0.8935 | 0.1710 |
| *VAC14* | All | 0.01 | 0.2814 | 0.4390 | 0.3819 | -1.2446 |
| *VAC14* | All | 0.03 | 0.3986 | 1.0000 | 0.7653 | -0.3362 |
| *VAC14* | Coding | 0.01 | 0.3683 | 0.7708 | 0.6414 | 0.0051 |
| *VAC14* | Coding | 0.03 | 0.3683 | 0.8155 | 0.6616 | -0.0728 |

## Discussion

Using the largest PD genetic and genomic datasets available to date, we conducted a comprehensive genetic assessment of NBIA related variants and genes for a potential role in PD etiology. We identified 28 previously reported NBIA coding mutations in a total of seven genes including *ATP13A2, CP, FA2H, C19orf12, FTL, PANK2, and PLA2G6* with a higher frequency in PD cases than controls. All phenotypes previously linked to the reported variants in NBIA genes involve neurological disorders, adding to the conclusion of clinical overlap between neurological diseases and the strong link between brain iron accumulation and a number of neurodegenerative disorders. However, only four out of the 28 variants of interest we reported had a statistically significant difference in frequency between cases and controls (p < 0.05), all only significant in UKB data. Within these four variants, one was found in *ATP13A2*, one in *FA2H*, one in *C19orf12*, and one in *PANK2*, always in only one case and no controls. Notably, none of these four variants have been previously associated with PD in the literature. Noting the very low mutation frequency in the current data and the lack of replication, we cannot conclude that the mutation spectrum contributing to NBIA phenotype overlaps with PD etiology. As such, despite enrichment for the risk allele in PD cases versus controls in four variants, these data suggest that in their heterozygous state NBIA variants are not frequently associated with risk of PD.

We performed burden analyses at two different frequency levels, MAF < 0.01 and MAF < 0.03, on both individual genes and as a gene-set. Meta-analysis of SKAT-O results were not significant for any individual gene or as a gene-set at either frequency level, suggesting that a cumulative effect of rare variation on NBIA genes does not play a role on PD etiology.

Transcriptomic analyses revealed that *DCAF17* is the only gene that is differentially expressed in blood from PD cases and controls. We further investigated this gene through Summary data-based Mendelian Randomization. One *DCAF17* eQTL signal was significant for both GWAS and SMR. The *DCAF17* gene is found on chromosome 2, encodes DDB1- and CUL4-associated factor 17, and is considered the cause of Woodhouse-Sakati syndrome, a rare neuroendocrine disorder which presents with neurological problems such as seizures and dystonia. ^66^ However, it is still unclear what is driving the significant *DCAF17* differential gene expression in our data and further assessments are needed.

This study has some limitations, mostly related to variant detection. Despite using the most well-powered PD datasets, it is nevertheless difficult to detect rare variants as evidenced by the number of variant counts of zero in both cases and controls. It is possible that other rare variants exist in NBIA genes that were not detected in our study. Therefore we cannot fully elucidate the relationship between rare variants in NBIA genes and PD. Additionally, we are only studying variants in individuals of European ancestry and could therefore be failing to detect disease-related genetic variation in other populations. A number of the variants we identified have been reported in the literature in non-European populations, so a study encompassing a greater genetic diversity would allow us to better characterize the relationship between NBIA and PD etiologies. Additionally, as we have mainly focused on coding variants, it is also possible that non-detected variants, such as larger structural variant (SVs) events in NBIA genes, affect PD risk. There is increasing evidence that SVs are likely the main contribution to disease susceptibility and have a substantial effect in coding regions of the genome where they can result in deleterious alterations of the DNA sequence.^67^ Therefore, further study into SVs would be an important step in identifying the relationship between NBIA and PD. Lastly, the cohorts currently available do not allow us to accurately perform an exhaustive clinical characterization of potential atypical symptoms, therefore possibly drawing limited conclusions. Additionally, as it has been previously suggested elevated nigral iron levels are unlikely to contribute to PD etiology but may vary with anti-parkinsonian drugs used for treatment.^68^ Taking everything into account, we suggest that even though NBIA and PD share similar symptoms, they could be molecularly different entities supporting the notion that the mechanisms underpinning iron accumulation in PD are not shared with NBIA. Elevated nigral iron levels may not contribute to PD etiology and may vary with anti-parkinsonian drugs used for treatment or any other environmental factors.

## Data Availability

All data used in the present study are available upon request from AMP-PD and UKB. Additionally, scripts and data produced are contained in the manuscript or available upon request to the authors.

https://github.com/neurogenetics/NBIA_PD

## Acknowledgements

Data used in the preparation of this article were obtained from Global Parkinson’s Genetics Program (GP2). GP2 is funded by the Aligning Science Against Parkinson’s (ASAP) initiative and implemented by The Michael J. Fox Foundation for Parkinson’s Research (**https://gp2.org**). For a complete list of GP2 members see **https://gp2.org**. The work was carried out with the support and guidance of the ‘GP2 Trainee Network’. This work was supported by the Center for Alzheimer’s and Related Dementias, within the Intramural Research Program of the National Institute on Aging and the National Institute of Neurological Disorders and Stroke, National Institutes of Health, Department of Health and Human Services. Project number 1ZIA AG000534-04.

C. Ruz held a predoctoral fellowship (FPU14/03473, MECD, Spain) from the Spanish Ministry of Education and Science.

**AMP-PD:** Data used in the preparation of this article were partly obtained from the Accelerating Medicine Partnership® (AMP®) Parkinson’s Disease (AMP PD) Knowledge Platform. For up-to-date information on the study, visit https://www.amp-pd.org. The AMP® PD program is a public-private partnership managed by the Foundation for the National Institutes of Health and funded by the National Institute of Neurological Disorders and Stroke (NINDS) in partnership with the Aligning Science Across Parkinson’s (ASAP) initiative; Celgene Corporation, a subsidiary of Bristol-Myers Squibb Company; GlaxoSmithKline plc (GSK); The Michael J. Fox Foundation for Parkinson’s Research ; Pfizer Inc.; Sanofi US Services Inc.; and Verily Life Sciences. ACCELERATING MEDICINES PARTNERSHIP and AMP are registered service marks of the U.S. Department of Health and Human Services. Clinical data and biosamples used in preparation of this article were obtained from the Michael J. Fox Foundation (MJFF) and National Institutes of Neurological Disorders and Stroke (NINDS) BioFIND study, Harvard Biomarkers Study (HBS), the NINDS Parkinson’s disease Biomarkers Program (PDBP) and MJFF Parkinson’s Progression Marker Initiative (PPMI). BioFIND is sponsored by The Michael J. Fox Foundation for Parkinson’s Research (MJFF) with support from the National Institute for Neurological Disorders and Stroke (NINDS). The BioFIND Investigators have not participated in reviewing the data analysis or content of the manuscript. For up-to-date information on the study, visit michaeljfox.org/news/biofind. The Harvard NeuroDiscovery Biomarker Study (HBS) is a collaboration of HBS investigators [full list of HBS investigator found at https://www.bwhparkinsoncenter.org/biobank/ and funded through philanthropy and NIH and Non-NIH funding sources. The HBS Investigators have not participated in reviewing the data analysis or content of the manuscript. PPMI – a public-private partnership – is funded by the Michael J. Fox Foundation for Parkinson’s Research and funding partners, a full list of the PPMI funding partners can be found at www.ppmi-info.org/fundingpartners. The PPMI Investigators have not participated in reviewing the data analysis or content of the manuscript. For up-to-date information on the study, visit www.ppmi-info.org. Parkinson’s Disease Biomarker Program (PDBP) consortium is supported by the National Institute of Neurological Disorders and Stroke (NINDS) at the National Institutes of Health. A full list of PDBP investigators can be found at https://pdbp.ninds.nih.gov/policy. The PDBP Investigators have not participated in reviewing the data analysis or content of the manuscript.

**UKB:** UK Biobank is a large-scale biomedical database and research resource containing genetic, lifestyle and health information from half a million UK participants. UK Biobank’s database, which includes blood samples, heart and brain scans and genetic data of the 500,000 volunteer participants, is globally accessible to approved researchers who are undertaking health-related research that’s in the public interest. UK Biobank recruited 500,000 people aged between 40-69 years in 2006-2010 from across the UK. With their consent, they provided detailed information about their lifestyle, physical measures and had blood, urine and saliva samples collected and stored for future analysis. UK Biobank’s research resource is a major contributor in the advancement of modern medicine and treatment, enabling better understanding of the prevention, diagnosis and treatment of a wide range of serious and life-threatening illnesses – including cancer, heart diseases and stroke. UK Biobank is generously supported by its founding funders the Wellcome Trust and UK Medical Research Council, as well as the Department of Health, Scottish Government, the Northwest Regional Development Agency, British Heart Foundation and Cancer Research UK. The organization has over 150 dedicated members of staff, based in multiple locations across the UK.

**23andme:** We thank the 23andMe research participants who consented to participate in research for enabling this study. We also thank employees of 23andMe.

## Author contributions

Initial manuscript preparation: Pilar Alvarez Jerez, Jose Luis Alcantud, Lucia de los Reyes, Anni Moore, Monica Diez-Fairen, Clara Ruz, Sara Bandres-Ciga, Raquel Duran.

Manuscript editing and commentary: Prabhjyot Saini, Ziv Gan-Or, Sara Bandres-Ciga, Francisco Vives Montero, Alastair Noyce, Andrew Singleton

## Ethical Compliance Statement

We confirm that we have read the Journal’s position on issues involved in ethical publication and affirm that this work is consistent with those guidelines.

## Code availabilityxs

Code can be found here: https://github.com/neurogenetics/NBIA_PD

