## Supplemental Table 1 for "Exploring the Genetic and Genomic Connection Underlying Neurodegeneration with Brain Iron Accumulation and the Risk for Parkinson’s Disease"

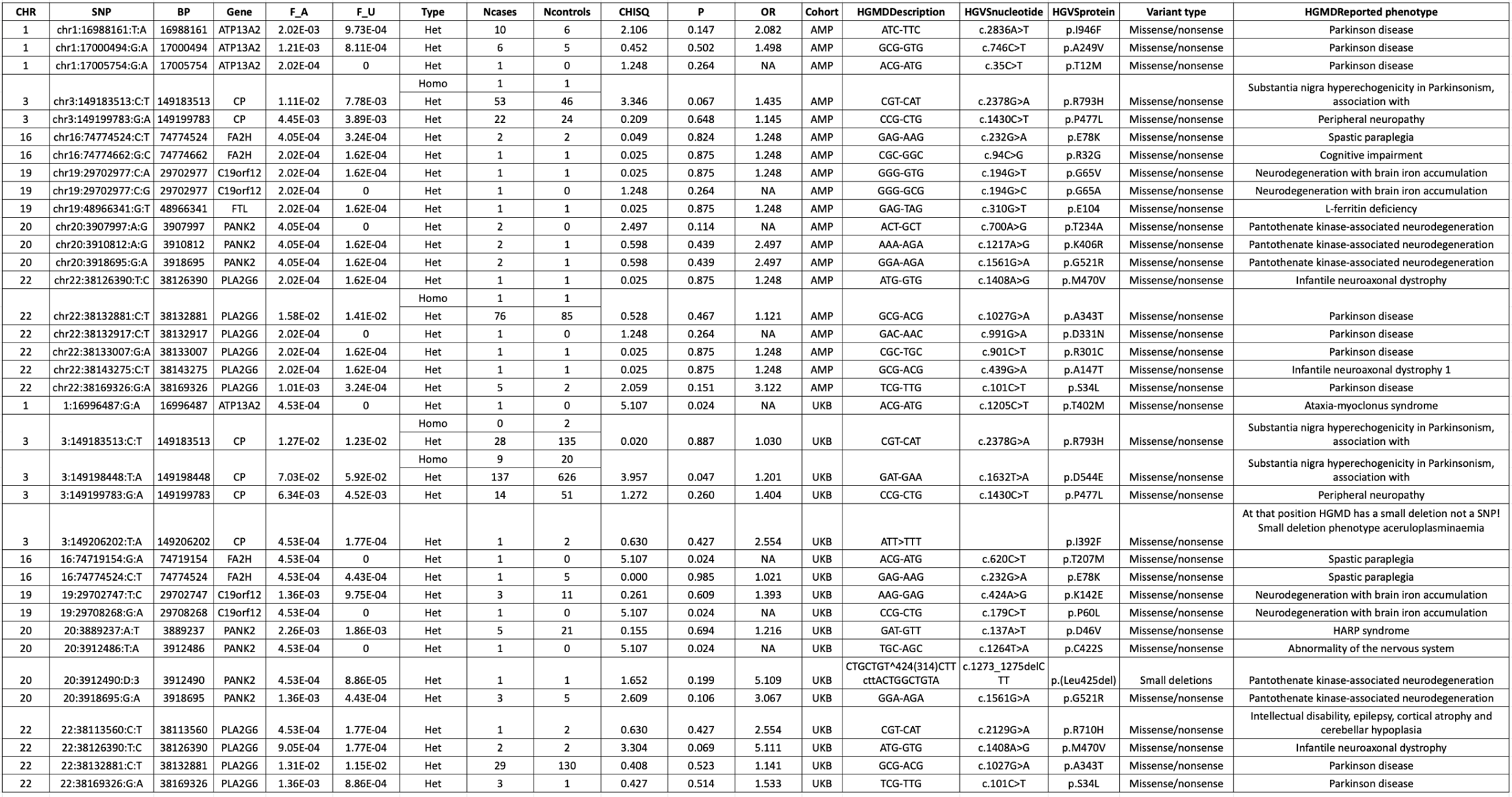

**Supplementary Table 1**: Screening of variants in NBIA related genes with higher frequency in PD cases versus controls. CHR = Chromosome, SNP=Single Nucleotide Polymorphism, BP= Base Pair, F_A= Frequency Affected, F_U= Frequency Unaffected, Ncases= Number of cases, Ncontrols= Number of controls, CHISQ= Chi-squared, P= P value, OR= Odds Ratio

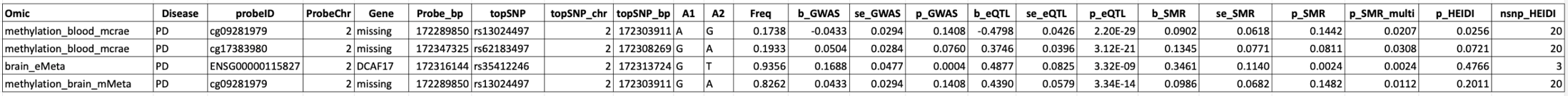

**Supplementary Table 2:** SMR results for *DCAF17*. Chr= Chromosome, BP= Base Pair, A1= Major Allele, A2= Minor Allele, b= Beta, se= Standard Error, p = p-value
